# Increased occurrence of migraine aura and spreading depolarizations at altitude

**DOI:** 10.1101/2025.08.09.25333153

**Authors:** Melissa M. Cortez, Katelyn M. Reinhart, Cecilia Martindale, Leon S. Moskatel, J. Gary Urry, Zubair A. Ahmed, Anna Newman, Kendra Pham, Jared Bartell, Todd Schwedt, Sheena Aurora, Kathleen B. Digre, Susan K. Baggaley, K.C. Brennan

**Author notes:** **Corresponding Author:** K.C. Brennan, **Corresponding Author’s Address:** 383 Colorow Drive Room 364, Salt Lake City UT 84108., **Corresponding Author’s., **Corresponding Author’s Email:**. These authors contributed equally to this work (Melissa M. Cortez, Katelyn M. Reinhart, Cecilia Martindale).

## Abstract

**Background and Objectives:** Test the hypothesis that migraine aura is more prevalent at altitude.

**Methods:** This report includes iterative examination of the association of aura and altitude using multiple studies derived from varying sources, including cross-sectional chart reviews, a cross-sectional multicenter registry study, a cross-sectional cross-institutional electronic medical record database study, and altitude chamber experiments in mice.

**Results:** We identify an association between increasing elevation and the presence of migraine aura. All four lines of clinical evidence converge on an altitude phenotype, with higher elevations being associated with higher odds of clinically defined aura. In data from mice housed in altitude chambers, we show that susceptibility to spreading depolarization (SD) is increased with simulated elevation, providing a candidate mechanism for the altitude phenotype.

**Discussion:** The increased susceptibility to aura and SD with elevation may provide a unique opportunity to develop targeted strategies for both research and clinical mitigation, relevant to the nearly 1 billion people living at higher elevations worldwide.

## Introduction

Migraine is a highly prevalent neurovascular disorder that affects 14-15% of the general population,^1,2^ and accounts for nearly 5% of all years lived with disability in the global population.^3^ Approximately 30% of those with migraine experience aura, which can impair quality of life by impacting driving, work, and caregiving responsibilities and is associated with higher migraine-related disability scores.^4,5^ More than 90% of migraine with aura patients experience transient visual disturbances, 36% experience somatosensory symptoms, and 10% experience language symptoms.^4,6^ In most, the migraine aura is the first clinically appreciable feature of the migraine (with aura) attack, and the underlying mechanism of aura, spreading depolarization (SD; also known as cortical spreading depression) is physiologically measurable in both animal models and humans. As a result, we have a growing understanding of how migraine with aura can lead to head pain.^7,8^ However, the precise mechanism(s) by which a massive depolarization like SD can be triggered in an apparently uninjured brain remain unclear.^7,8^ Critically, having a known physiological mechanism in SD^7,8^ allows us to more explicitly examine candidate mechanisms underlying the relationship between aura and altitude.

Prior work has suggested that both migraine and migraine aura prevalence may be greater at higher altitudes. Multiple potential mechanisms have been proposed, including hypoxia,^9,10^ barometric pressure differences,^11,12^ increased solar radiation/sunlight,^13,14^ metabolic challenge,^15,16^ dehydration,^17^ and a pro-inflammatory environment.^18,19^ However, even the largest human studies, have not explicitly examined how the presence of migraine associated aura interacts with altitude^20,21^. The objective of this report is to test the hypothesis that aura, measured clinically (human studies) and physiologically (animal model), is more common with increasing elevation. Here we summarize the findings of multiple lines of data that examine the relationship of migraine aura and altitude. The series of studies began with an anecdotal clinical insight, namely that there appeared to be a greater prevalence of migraine aura in a headache clinic at moderate (∼1400m) elevation. We subsequently sought to test this observation through a series of studies spanning multiple clinical settings, each building on the last toward increasingly broader data sources, to evaluate whether the initial observation was indeed generalizable. Finally, we hypothesis-tested the potential mechanism of our clinical data in an animal model.

## Materials and methods

### Study 1

Study 1 was a retrospective cross-sectional investigation of the proportion of patients with migraine presenting with aura at two different sites. We reviewed the electronic medical records (EMR) of new, consecutive patient visits presenting to headache specialty clinics between January and October 2011. Clinic sites represented moderate and low elevations: Salt Lake City (SLC), Utah (University of Utah Headache Clinic, elevation 1400m; 3 providers, 272 patients) and Seattle, Washington (Swedish Headache Center, elevation 57m; 1 provider, 199 patients). The study protocol was approved by the Institutional Review Boards from both the University of Utah and the Swedish Headache Center.

Eligibility criteria included any patient presenting for new evaluation by a headache specialist at the center, and a diagnosis of migraine with or without aura at the initial encounter, as documented in their medical record. There were no explicit exclusion criteria outside of the stated eligibility requirements. Non-migraine headache was not included, while mixed headache types were allowed, though not explicitly coded outside of the migraine classification. Migraine with and without aura were classified through clinical history and examination, assigned, and documented by providers according to the International Classification of Headache Disorders (ICHD) criteria in effect during the study period (ICHD-II). Our outcome of interest was the proportion of documented migraine aura within the sampled patients with migraine diagnosis. Additionally, sex and age were collected from the EMR; body mass index [BMI] was also recorded to evaluate for site-dependent variations in clinical population.

We compared migraine with aura diagnosis between the low and moderate elevation groups (Pearson’s chi-squared test for association), as well as other clinical variables (two-sample t-test/Kruskal-Wallis or chi-square tests as appropriate); logistic regression was used to adjust for age and sex (statistical significance evaluated using the Wald test). Results are reported as odds ratios, reflecting the change in odds of aura between the two sites given that, unlike the subsequent studies (Studies 2-4), Study 1 did not feature specific elevations. Thus, while the analyses across Studies 1-4 are related, odds ratios from Study 1 are not directly comparable to odds ratios from Studies 2 and 3 that reflect the change in odds of aura with elevation.

### Study 2

Study 2 was a prospective cross-sectional investigation of migraine aura at two clinical sites. This second study differed from Study 1 in participant selection, using refined inclusion/exclusion criteria, inclusion of residential-level altitude for each participant, and an expanded characterization of aura using the Visual Aura Rating Scale (VARS). VARS was selected as a well-validated instrument with excellent sensitivity and specificity (96.4% and 79.5%, respectively) for the diagnosis of migraine-associated aura.^22^ VARS criteria include aura duration between 5-60 minutes, presence of scotoma, zigzag lines, unilateral visual field involvement, and visual symptoms developing gradually over at least 5 minutes. A cut-off score of 5 or more out of a maximum of 10 points has been shown to reliably classify migraine-related aura status, based on prior work.^23^ Aura status, as well as sex and age, were considered primary variables of interest. Additional clinical variables of ethnicity, caffeine intake, and BMI were also recorded to evaluate for site-dependent variations in clinical population.

Patients presenting to the University of Utah Headache Clinic (August 2015 to January 2016) and the Cleveland Clinic Headache Clinic (199m elevation; August 2016 - February 2017) were included. Eligibility status was determined by a single, dedicated clinician rater [ZA; trained in Headache Medicine, working consecutively in both locations through the period of the study]. Participant altitude of residence was coded using address-associated ZIP codes. To determine elevation of residence, we derived ZIP code level mean elevation from Shuttle Radar Topography Mission satellite image estimates, which were available via *ecolo-zip.*^24^ This approach enabled us to approximate participant residential elevations rather than relying on the elevation of participants’ presenting clinical site.

Inclusion criteria were age 18 years old or greater with a (Headache Specialist) clinician-assigned, chart-documented diagnosis of migraine. Exclusion criteria were the presence of other, non-migraine neurological conditions, disorders associated with hypercoagulability, and/or inability to provide informed consent. As in Study 1, individuals with non-migraine headache were not included, though those with mixed headache types (e.g., migraine and tension headache) were allowed, though not explicitly coded outside of the migraine classification.

For analysis purposes in Study 2, our outcome of interest was aura (coded as a dichotomous variable, yes/no, based on a VARS cut-off score of 5 or more^23^), and our exposure of interest was elevation of residence. We compared aura percentages across the two sites (Pearson’s chi-squared test for association), as well as other clinical variables (Kruskal-Wallis tests or chi-squared as appropriate). We then assessed the association between elevation and odds of aura, adjusting for the effects of age and sex, using a multivariable logistic regression model (statistical significance evaluated via the Wald test). Results are reported as odds ratios, reflecting the change in odds of aura with each 100m difference in altitude.

### Study 3

In Study 3, we examined aura and elevation data using data from a national database of headache specialty clinics, the American Registry for Migraine Research (ARMR). The initial ARMR study, described in detail elsewhere,^25^ enrolled patients with headache disorders from 2016 to 2020 at eight participating health centers (University of Utah, University of Colorado, Mayo Clinic Arizona, Dartmouth, Dent Neurologic Institute, Georgetown, University of Texas, and Thomas Jefferson University) and collected comprehensive patient-reported data on headaches and related symptoms, including the VARS. For consistency of methodology across Studies 2 and 3, the presence and severity of aura were determined using the VARS to code participant aura grouping (yes/no aura), following the same criteria outlined in Study 2.

For our secondary analysis of these data, inclusion criteria included participants for whom an ICHD-3 diagnosis of “migraine” (any) had been assigned, and where sex, age and ZIP code data were available; we excluded duplicates and those missing the patient-reported headache feature questionnaire.^25,26^ Of the eight participating ARMR sites, three provided ZIP code information. To determine elevation of residence, ZIP code level mean elevation was derived via satellite estimates as described above in Study 2.^24^ In addition to ZIP code of residence reported at baseline during enrollment, we also extracted VARS, age and sex as primary variables of interest; race (white or non-white), depression score (PHQ-4), MIDAS, and headache days per month were also extracted to evaluate for site-dependent variations in clinical population.

For this analysis, our designated outcome was migraine aura, and the exposure of interest was elevation of residence. We compared aura prevalence across the included ARMR sites (Pearson’s chi-squared test for association), as well as other clinical variables to evaluate site-specific differences (t-test, Kruskal-Wallis tests or chi-squared as appropriate). We assessed the association of elevation of individual-participant residence with VARS-defined aura prevalence using logistic regression, with elevation as the primary predictor and aura (dichotomous, yes/no) as the response variable, adjusted for age and sex. Results are reported as odds ratios, reflecting the change in odds of aura with each 100m difference in altitude.

Finally, to better understand how elevation was associated with aura-specific factors, we examined relationships between elevation and specific VARS items -- 1) aura onset, 2) duration, 3) scotoma, 4) zigzag lines, and 5) unilateral location -- using logistic regression and adjusting for age and sex. These aura-specific factors were only available for Study 3. Statistical significance was assessed using a Wald test. We adjusted p-values for multiple comparisons using Holm’s correction.

### Study 4

To further evaluate the relationship of migraine aura to elevation of residence at a national level, without the site-level biases of our prior work, we utilized the Epic Cosmos Research Platform (Epic System Corporation, Verona, WI) to evaluate nationwide county-level aura prevalence by elevation. The Epic Cosmos Research Platform is an aggregated, de-identified database of electronic medical records (EMR) from health systems across the United States, specifically designed for research purposes, and consists of over 300 million patient records from over 1,744 hospitals and 40,700 clinics from all 50 states and three other nations; only patients in the United States were included in this study.^27^ Of note, Epic Cosmos data is known to generally reflect the United States population’s demographics.^28^ As individual-level ZIP code data was not available with our access to the database, we aimed to estimate elevation of residence via county-level designations using the standard digital elevation model from the United States Geological Survey (USGS) elevation dataset.^29^ This database provides latitude and longitude coordinates for each county seat and returns the corresponding elevation value for any location within the United States. To ensure accuracy, county seat elevation data was verified using Google Earth elevation data.

To obtain our primary sample, we queried the Cosmos database for patients in the United States with an International Classification of Diseases, Tenth Edition (ICD-10) code of G43 “Migraine,” or higher resolution, over the five-year period of January 2020 - December 2024. Patients were classified as having aura based on the presence of an ICD-10 code of G43.1, indicating ‘Migraine with aura’ (or a more specific diagnosis) during this same time frame. Epic Cosmos-based ICD diagnoses were aggregated to the county level, based on patient address of residence as recorded in the EMR. We then calculated the percentage of patients with a migraine diagnosis who also had codes for migraine with aura within each county. Of note, this data gives an EMR prevalence that may or may not be generalizable to the population at large. Alaska was excluded due to insufficient number of patients and the District of Columbia was not included as it is a single county. Counties without Epic Coverage could not be included. Counties with fewer than 10 patients with migraine with aura during the queried time period were excluded as Epic Cosmos does not provide exact numbers of patients for data elements fewer than 10 patients.

For this analysis, we performed ordinary least squares-based linear regression using the percentage of patients with migraine who had migraine with aura at the county level as the dependent variable and the county seat altitude (in feet) as the independent variable, adjusted for age, sex, and state. State-effects were controlled for through clustering at the state level to account for repeat measurements in each state. The results of these linear regression models are presented as the coefficients measuring the percentage point change in aura prevalence among Epic Cosmos patients with migraine at the county level for each increase in elevation. Note that these results thus differ from the odds ratios presented for Studies 2 and 3, as noted above.

### Mouse studies

All procedures were conducted in accordance with protocols approved by the University of Utah Health Sciences Center Institutional Animal Care and Use Committee. Experiments used adult male mice (C57Bl/6J, 1.5 – 4 months old, 22.6 ± 0.2g, Jackson Laboratory, Bar Harbor ME), housed in a temperature and humidity-controlled vivarium with a 12-hour light/dark cycle and free access to food and water. Detailed mouse study methods are provided in the Supplemental Methods.

#### Altitude exposure

We employed custom-made altitude chambers to test the effect of high (4500m, 14760ft), moderate (1400m, 4594ft; lab altitude in SLC), and low (0m) altitudes on SD threshold in mice. Aluminum vacuum chambers (5 gallon, 1931N18 McMaster-Carr, Robbinsville, NJ, USA) were fitted with adjustable pressure-relief valves and connected to vacuum (high altitude) or pressure (low altitude) pumps to maintain the desired atmospheric pressure within the chamber (101kPa for 0m, 86kPa for 1400m, and 58kPa for 4500m^30^). Pre-calibrated sensors (NeuLog, Rochester, NY, USA) continuously monitored absolute pressure (in kPa; NUL-210 sensor) and effective oxygen concentration (in %; NUL-205 sensor). Following 2–5 days of habituation to the chambers (4 – 5 mice/chamber; mice were maintained at local altitude pressures but acclimated to the noise of vacuum and pressure pump operation) pressure-relief valves were incrementally adjusted at a rate of ≤1 kPa/min. Once target altitude pressures were reached, chamber conditions were maintained for 10 days. On day ten, mice were returned to local lab conditions (≤1 kPa/min), and experiments were conducted that same day by an experimenter blinded to the altitude group being tested. An independent researcher, who was not involved in SD thresholding and counting experiments, monitored all chambers and interleaved altitude groups to maintain blinding.

#### Surgical preparation

Experiments were conducted in each mouse under isoflurane anesthesia. For SD thresholding experiments, two burr holes (0.5mm diameter) were made in the right hemisphere for SD initiation (AP: +3.0mm ML: +1.0mm) and electrophysiological detection (AP: - 3.0mm, ML: +3.0mm). For SD counting experiments, a 1.5mm diameter craniotomy was created in the right barrel cortex (AP: −1.5mm ML: +2.5mm) to expose the brain for SD induction. Additional details are provided in Supplemental Materials.

#### SD thresholding

SD threshold was evaluated using our previously published methods.^31^ Briefly, we used a borosilicate glass pipette (22-24μm) connected to a pneumatic pico-pump to stepwise increase the volume of 1M KCl delivered focally into a cranial burr hole until SD was initiated. The volume of KCl was increased every five minutes using 20ms square-wave pressure pulses (from 2.5 psi up to 15 psi) and SD was detected using a local field potential (LFP) electrode placed a second distal burr hole. SD was confirmed via its large characteristic direct current shift (DC shift) in the extracellular slow potential.

#### SD counting

SD counting procedures were performed using our previously published methods^31^. Briefly, 1M KCl was perfused at 0.016mL/min over the SD initiation site for a total of 2 hours. SD numbers were then totaled over the 2-hour period for each animal.

#### Hematocrit determination

After SD experiments, blood was collected from the tail of each mouse into 60mm heparinized microhematocrit tubes (VWR International, Randor, PA, USA). Capillary tubes were centrifuged at 13,000 x *g* for 3 min and the Hct value (%) was measured according to the DIN 58933 practical standard reading graph.^32^

### Statistics, Reporting of Data, Rigor

Analyses of the clinical data used R version 4.3.3 and RStudio Version 1.4.1717 or Stata 14. For descriptive statistics, categorical variables are presented as count with percent, normally distributed continuous variables are presented as mean with standard deviation, and skewed continuous variables are presented as median with interquartile range. Descriptive statistics are presented in aggregate and stratified by group. We followed the STROBE and ARRIVE checklists for human and animal data, respectively.

Analysis of the animal data used Graphpad Prism 10.1.1 and each animal served as an individual experimental unit. The experimenter remained blinded to altitude group assignments until completion of all experiments and primary data analyses, after which group identities were revealed for statistical analyses (details provided in Supplemental Methods).

### Data Availability

Anonymized data not published within this article will be made available by request from any qualified investigator.

## Results

### Study 1

Demographic data and aura prevalence for Study 1 are summarized in **Table 1**. Of the total sample of 471 participants, 74.3% were female (*n* = 350), with a median age of 42.0 years (range 18-91; IQR = 20.0). There were no significant differences between sites for age, sex, or BMI.

The proportion of participants with migraine with aura was significantly higher in those presenting for care in SLC (site at 1400m; moderate elevation) compared to those presenting in Seattle (57 m), with percentages of 53.3% and 17.6%, respectively (*P* < 0.0001). Participants in Seattle had significantly lower odds of aura compared to those in SLC after adjusting for age and sex (OR 0.18, 95% CI: 0.12, 0.28), corresponding to approximately 5.5-fold higher odds of aura among SLC participants compared to those in Seattle. Additional details of the regression model outputs are summarized in Supplemental Table S1.

**Table 1.** Demographic features and aura prevalence by site for Study 1.

|  | **Total**<br>n=471 | **Seattle**<br>n=199 | **Salt Lake City**<br>n=272 | **P value** |
| --- | --- | --- | --- | --- |
| **Age**, median (IQR);<br>minimum-maximum | 42.0 (20.0);<br>18-91 | 43.0 (19.0);<br>19-64 | 40.0 (22.3);<br>18-91 | 0.379 |
| **Female**, n (%) | 350 (74.3) | 157 (78.9) | 193 (71.0) | 0.051 |
| **BMI**, mean (StDev);<br>min-max | 28.7 (7.3);<br>17-54 | 28.3 (7.1);<br>17-54 | 29.2 (7.6);<br>18-53 | 0.271 |
| BMI category, n (%) |  |  |  |  |
| Underweight | 6 (1.3) | 5 (2.5) | 1 (0.4) |  |
| Healthy weight | 108 (22.9) | 72 (36.2) | 36 (13.2) |  |
| Overweight | 92 (19.5) | 57 (28.6) | 35 (12.9) |  |
| Obese | 110 (23.4) | 64 (32.2) | 46 (16.9) |  |
| Missing | 155 (32.9) | 1 (0.5) | 154 (56.6) |  |
| **Aura**, n (%) | 180 (38.2) | 35 (17.6) | 145 (53.3) | **< 0.001** |

### Study 2

Demographic data and aura prevalence for Study 2 are summarized in **Table 2**. Of the 208 participants studied, 86.1% were female (*n* = 179), with a median age of 40.0 years (range 18-76 years; IQR = 21.0). There were no significant differences between sites for sex or caffeine intake; whereas age was significantly higher in SLC compared to Cleveland, participants were more likely to be white in SLC, and BMI was higher in Cleveland.

**Table 2.** Demographic features and aura prevalence by site for Study 2.

|  | **Total**<br>n=208 | **Cleveland**<br>n=104 | **Salt Lake City**<br>n=104 | **P value** |
| --- | --- | --- | --- | --- |
| **Age**, median (IQR);<br>min-max | 40.0 (21.0);<br>18-76 | 35.0 (18.00);<br>19-72 | 44.5 (21.3);<br>18-76 | **< 0.001** |
| **Female**, n (%) | 179 (86.1) | 92 (88.5) | 87 (83.7) | 0.317 |
| **Race**, n (%) |  |  |  | **< 0.001** |
| White | 165 (79.3) | 73 (70.2) | 92 (88.5) |  |
| Non-white | 42 (20.2) | 31 (29.8) | 11 (10.6) |  |
| Missing/not reported | 1 (0.5) | 0 | 1 (1.0) |  |
| **BMI**, mean (StDev);<br>min-max | 27.9 (6.4);<br>18-53 | 29.2 (6.1); 19-53 | 26.9 (6.3); 18-48 | **0.004** |
| BMI category, n (%) |  |  | 3 (2.9) |  |
| Underweight | 3 (1.4) | 0 (0) | 42 (40.4) |  |
| Healthy weight | 59 (28.4) | 17 (16.2) | 28 (26.9) |  |
| Overweight | 50 (24.0) | 23 (21.9) | 31 (29.8) |  |
| Obese | 68 (32.7) | 37 (35.2) | 0 (0) |  |
| Missing | 28 (13.5) | 28 (26.7) |  |  |
| **Caffeine intake**, n (%) |  |  |  | 0.496 |
| None | 43 (20.7) | 21 (20.0) | 22 (21.2) |  |
| 12oz or less | 88 (42.3) | 49 (46.7) | 40 (38.5) |  |
| >12 oz | 75 (36.1) | 34 (32.4) | 41 (39.4) |  |
| Missing | 2 (1.0) | 1 (1.0) | 1 (1.0) |  |
| **Aura (VARS ≥ 5)**, n (%);<br>min-max | 89 (42.8); 0-10 | 35 (33.7);<br>2-10 | 54 (51.9); 0-10 | **0.008** |

The percentage of migraine with aura within the SLC-based group (moderate elevation) was significantly higher than in the Cleveland-based group (51.9% in SLC 1400m vs 33.7% in Cleveland 199m respectively; *P* = 0.008). Binary logistic regression showed that a difference of 100m elevation was associated with 3.7% increase in odds of aura (OR per 100m = 1.037, 95%CI: 0.997, 1.078). Thus, for example, when comparing individuals with migraine at 1000m of elevation to sea level this model estimates 43% higher odds of aura for individuals with migraine at 1000m, compared to those at sea level. Additional details of the regression model outputs are summarized in Supplemental Table S2.

### Study 3

Our initial sample from the ARMR national headache database was composed of data from three ARMR sites with available ZIP code data, yielding an initial total of 1016 participants; 71 were excluded due to lack of ZIP code data; 36 were excluded because they held a different primary ICHD diagnosis than migraine or no diagnosis assignment was included in the ARMR database; and 25 were excluded due to incomplete headache features data needed to calculate aura scores. Thus, our final sample included 884 migraine patients for analysis. **Figure 1** is a schematic of participant exclusions.

**Figure 1.**
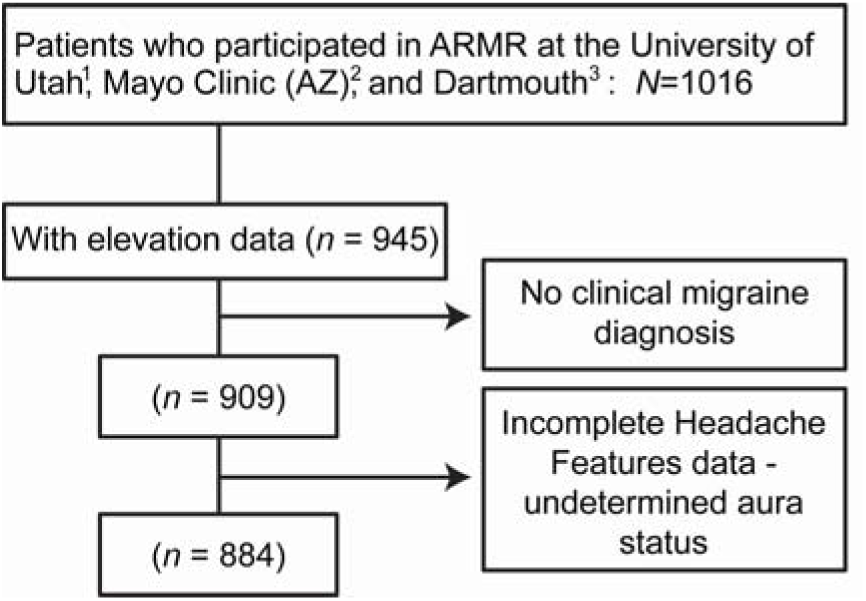
Participant exclusions in Study 3. 1: University of Utah, Salt Lake City, UT. 2: Mayo Clinic, Scottsdale, AZ. 3: Dartmouth-Hitchcock Medical Center, Hanover, NH.

Participant characteristics for Study 3 are summarized in **Table 3.** Of the 884 participants included in our analysis, 86.9% were female (*n* = 768), with a median age of 47 (range 18-86; IQR = 21.0). There were no significant differences between sites for sex, nor headache days/month, whereas age, ethnicity, PHQ-4, and MIDAS differed across sites. The mean elevation across the entire sample was 673m (range 0.305m – 3220m); number of participants living at each 100m elevation category are summarized in Supplemental Table S3. Note that the range of elevation of residence was greater than the local altitude range around each clinical site (216 – 1400m), likely reflecting that patients sometimes traveled long distances to these tertiary referral centers.

**Table 3.**
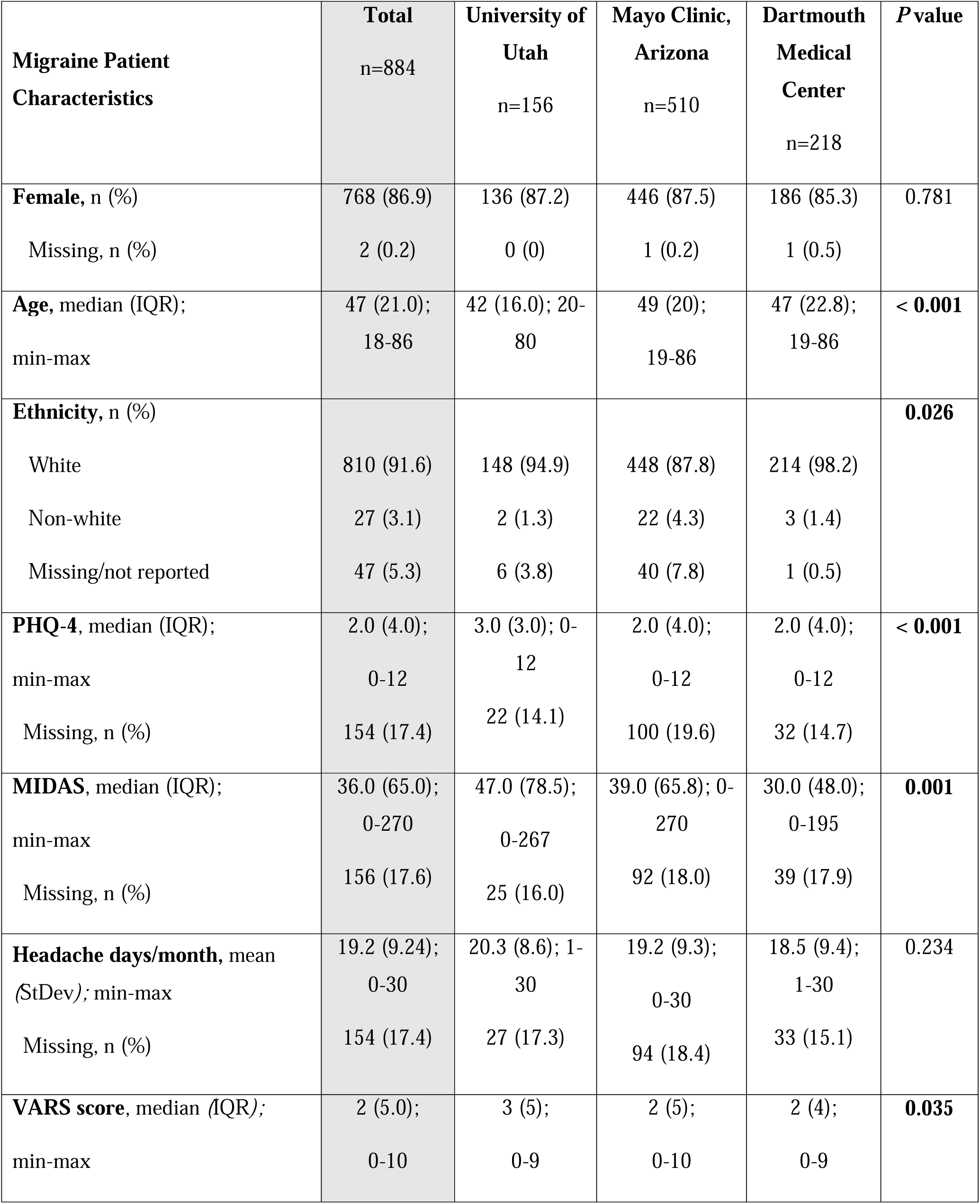
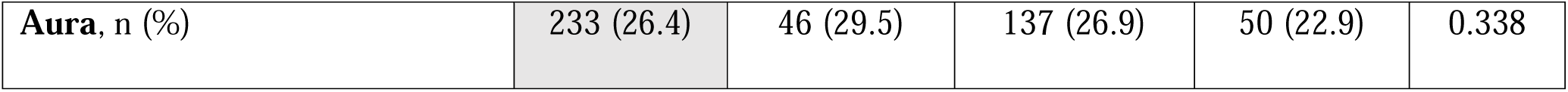
Demographic features and aura prevalence by site for Study 3.

Binary logistic regression analysis of elevation of residence and aura status (as a dichotomous yes/no), while adjusting for age and sex, across the full ARMR sample provided an odds ratio of (OR: 1.029; 95% CI: 1.003, 1.056). Thus, for each incremental increase of 100m (328ft), the odds of aura were 2.9% higher (95%CI: 0.3, 5.6%). For example, when comparing individuals with migraine at 1000m of elevation to sea level, this model estimates a 33% greater odds of aura among the individuals at 1000m. **Figure 2** plots aura prevalence by elevation for the ARMR cohort to illustrate this relationship. Results of regression analyses for Study 3 are summarized in Supplemental Table S4 (primary VARS-based analysis), including additional sensitivity analyses utilizing aura as a clinician-assigned diagnosis in Table S5.

**Figure 2.**
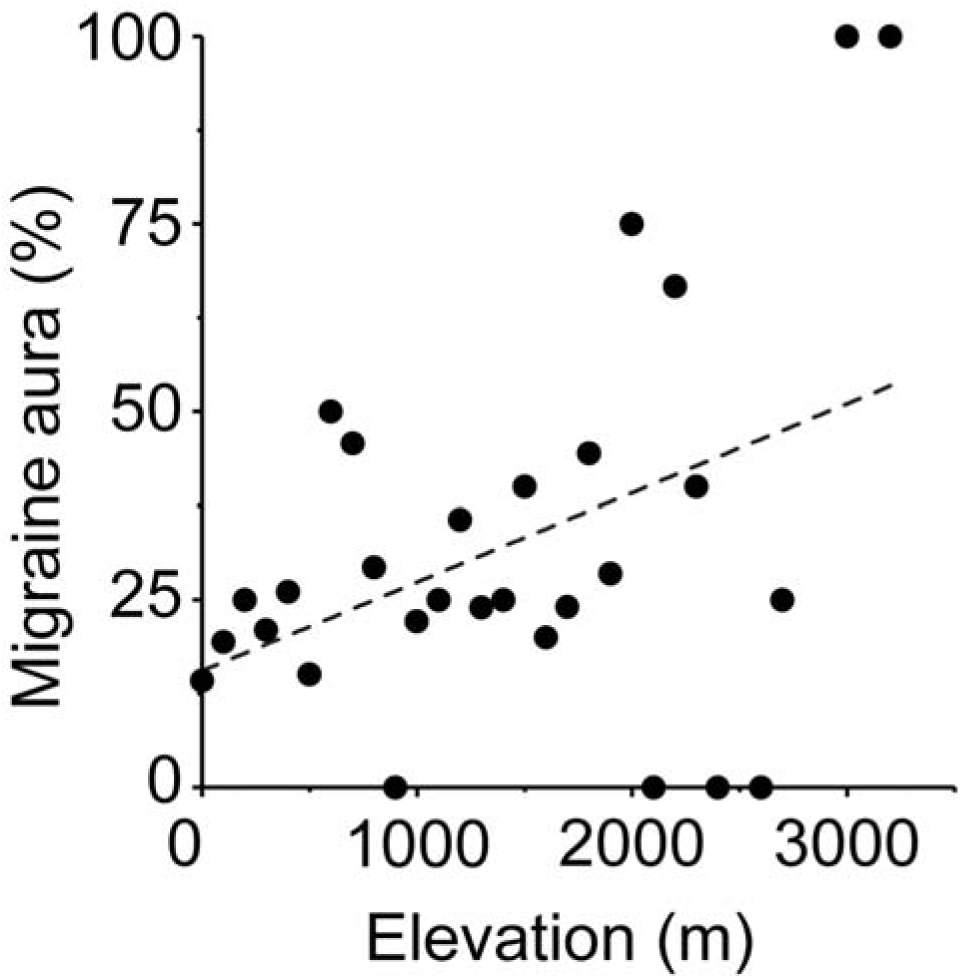
Aura percentage comparison against elevation in Study 3. We compared aura percentage by elevation in increments of 100m (328ft). Each point represents the prevalence of migraine aura among ARMR participants at that elevation. <u>Note</u>: the proportions at the highest elevations (> ∼1900m) reflect very small sample sizes and should thus be interpreted with caution. Detailed sample size (n) per elevation interval is available in the Supplemental Materials (Table S3).

Finally, logistic regression, examining the relationship of elevation and specific aura features via VARS (aura onset, duration, scotoma, zigzag lines, and unilateral location), and adjusting for age and sex, found a significant relationship between scotoma and elevation, where an increase in elevation of 100m was associated with 5.2% higher odds of scotoma (OR: 1.052; 95% CI: 1.024, 1.080). In contrast, the other VARS-based aura features of onset, duration, zigzags, and location were not significantly associated with elevation. Results of the VARS-feature based regression analyses are summarized in Supplemental Table S6.

### Study 4

The initial query of the Epic Cosmos database yielded county-level data for 3109 counties across 49 included states. A total of 2,685/3109 (86.4%) of these counties had at least 10 patients with migraine with aura, as well as average age and percentage of female patients available, and were included in the analysis. These counties had a median elevation of 259m (range −2 to 3221m; IQR 134, 4100) and an average migraine aura prevalence of 27.2±5.4% in this single EMR-based sample. **Figure 3** is a schematic of participant exclusions.

**Figure 3.**
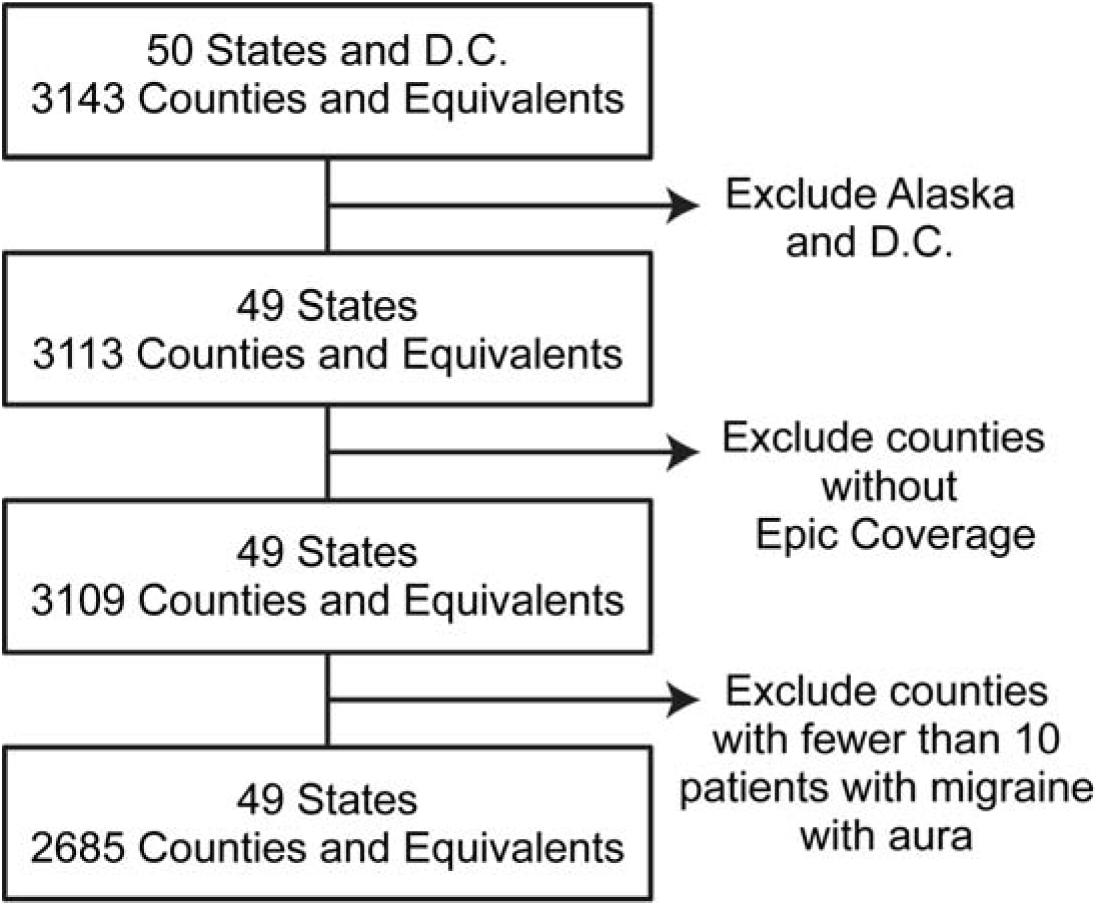
Participant exclusions in Epic Cosmos cohort.

Linear regression with ordinary least squares, adjusting for age, sex, and potential state-effect (which was controlled for with clustering at the state level), demonstrated a statistically significant association between county-level elevation and migraine aura prevalence in the EMR. With each 100m increase in elevation, we observed a 0.129 percentage point increase in migraine aura prevalence per 100m increase in elevation (95% CI: 0.014, 0.245). This suggests that patients at an elevation of 1000m would have a 1.29 percentage point higher prevalence of migraine aura relative to those living at sea level (**Figure 4**). Additional details of the regression model outputs are summarized in Supplemental Table S7.

**Figure 4.**
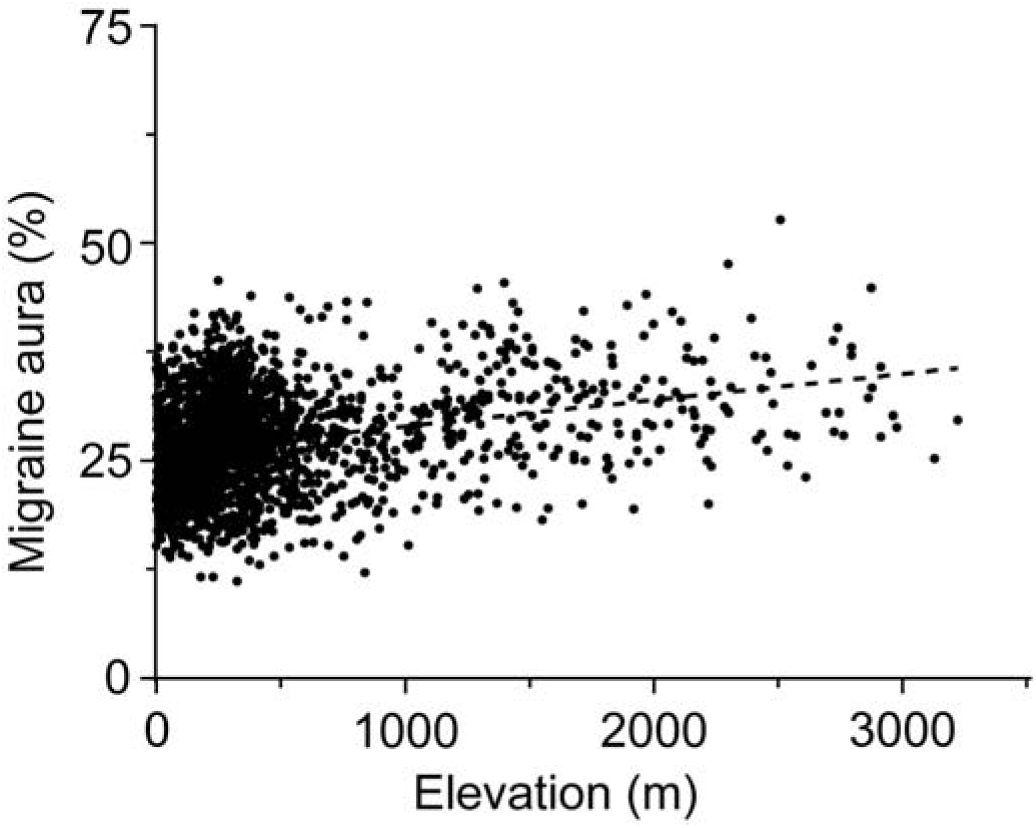
Aura percentage comparison against elevation in Study 4. We compared aura rate percentage by elevation for each county included in the Epic Cosmos cohort. Each point represents county-specific elevation and rate prevalence of migraine aura.

### Mouse Study

Although SD is widely recognized as the physiological mechanism underlying migraine aura,^7,8^ its presence is almost always inferred clinically as it is very difficult to measure (either invasively^33^ or non-invasively^34^) in migraine patients. We therefore tested in mice whether the increased prevalence of aura observed in patients living above sea level may be due to a greater susceptibility to SD. To test this, we exposed mice to high (4500m), moderate (1400m, local lab altitude), and low (0m, sea level) altitudes for 10 days (**Figure S2A)** and then used two methods to evaluate SD susceptibility. Throughout the experiment, atmospheric pressure (in kPa) and effective oxygen percentage were continuously monitored in each chamber and remained stable (**Figure S2B**).

We first evaluated the influence of altitude on the threshold at which SD could be evoked, using male C57Bl/6J mice. After altitude manipulations, SD threshold was examined using focal microinjection of 1M KCl,^31^ the volume of which was incrementally increased (via stepped pressure pulses every 5 min) until SD was detected at a distant recording electrode (**Figure 5A**). Results in **Figure 5B** show that SD threshold differed significantly across altitude groups (Kruskal–Wallis test, *P* < 0.0001). Moreover, SD was evoked using the lowest volume of KCl tested in the majority of high (14/22; 64%) compared to local (7/22; 32%) and sea level (1/22; 4.5%) mice. This corresponded to SD threshold being significantly lower in mice exposed to 4500m compared to both 0m (*P* < 0.0001) and 1400m (*P* = 0.015). Although the difference between 0m and 1400m did not reach statistical significance (*P* = 0.053), these differences corresponded to large effect sizes for comparisons involving the 4500m group (0m vs 4500m: Cohen’s *d* = 2.07, 95% CI [1.50, 3.06]; 1400m vs 4500m: *d* = 1.12, 95% CI [0.63, 1.76]) and a moderate effect size for the 0m vs 1400m comparison (*d* = 0.84, 95% CI [0.25, 1.58]). To further validate the SD susceptibility altitude phenotype, we employed repetitive SD induction, a second widely used SD evaluation.^31,35^ In these experiments, cortical intrinsic signal imaging was used to monitor SD frequency over a 2-hour recording period, with continuous KCl perfusion serving as the evoking stimulus (**Figure 5C**). As shown in **Figure 5D**, the number of SDs was significantly increased at 4500m (15.4 ± 1.0 SDs) compared to both 1400m (12.3 ± 0.7 SDs, *P* = 0.025) and 0m (10.8 ± 0.6 SDs, *P* = 0.001), indicating an altitude-related enhancement in SD occurrence. Similar to SD thresholding results, differences were driven primarily by contrasts involving the 4500m group, which showed large effect sizes (0m vs 4500m: Cohen’s *d* = 1.43, 95% CI [0.80, 2.44]; 1400m vs 4500m: *d* = 0.96, 95% CI [0.28, 1.91]), while the comparison between 0m and 1400m demonstrated a graded but non-significant progression (*d* = 0.61, 95% CI [−0.12, 1.43]).

**Figure 5.**
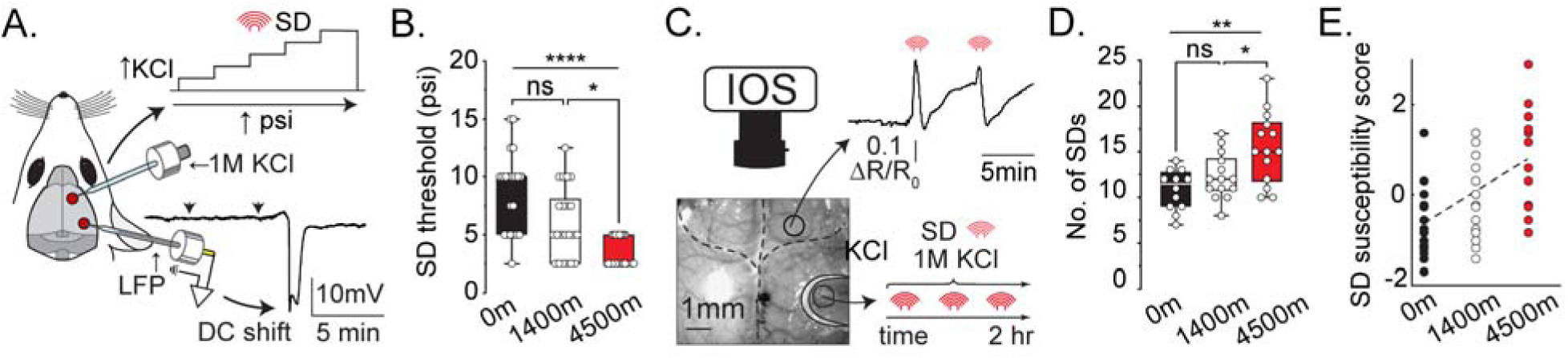
High altitude lowers SD threshold and increases SD susceptibility in mice. **A.** Schematic showing the location of burr holes utilized for SD induction (rostral; for potassium chloride [KCl] administration) and detection (caudal location; for recording of local field potentials [LFP]). Stepped increases in pressure (psi) were applied to the micropipette at 5-minute intervals to deliver increasing volumes of KCl into the rostral location until SD was detected at the distant LFP electrode (see representative direct current [DC] shift trace). Arrowheads indicate time points of KCl application. **B.** Group data shows significantly lower SD thresholds in C57Bl/6J mice exposed to 4500m compared to those in 1400m and 0m altitude groups (*n* = 22 mice/group). **** *P* < 0.0001, * *P* = 0.015, n.s. *P* = 0.053; Kruskal-Wallis with Dunn’s post hoc multiple comparisons test. **C.** Schematic of the experimental design for repetitive SD induction using intrinsic optical imaging (IOS) of the dorsal surface of the skull (see representative image and reflectance trace during two SDs). KCl was slowly perfused onto the exposed cortex of the right hemisphere for 2 hours. **D.** Group data show a significantly higher number of SDs detected over the testing period in mice exposed to 4500m compared to both 0m (*n* = 12) and 1400m (*n* = 14) altitude groups (*n* = 14 at 4500m). * *P* = 0.025, ** *P* = 0.001, n.s. *P* = 0.67; one-way ANOVA with Bonferroni’s post hoc multiple comparisons test. **E.** Composite SD susceptibility score generated via post hoc z-score normalization of SD thresholding and SD counting data shown in panels B and D (*n* = 34, 36, 36 for 0, 1400, and 4500m groups, respectively). Linear regression analysis demonstrated a significant positive association between altitude and SD susceptibility score (Y = (3.12 × 10□□) X - 0.625, R^2^ = 0.350, *P* < 0.0001), with a slope of 3.12 × 10□□ (95% CI: 2.29 × 10 □□to 3.95 × 10□□), consistent with a graded increase in SD susceptibility across altitudes.

To aggregate SD susceptibility across both assays, we generated a post hoc composite “SD susceptibility score” using z-score normalization (**Figure 5E**, and Supplemental Methods). Pairwise comparisons of the composite SD susceptibility score showed moderate to large effect sizes across altitude contrasts (0m vs 1400m: Cohen’s *d* = 0.74, 95% CI [0.29, 1.22]; 0m vs 4500m: *d* = 1.84, 95% CI [1.32, 2.58]; 1400m vs 4500m: *d* = 0.99, 95% CI [0.52, 1.59]), consistent with the graded pattern observed in the individual SD thresholding and SD counting assays. Linear regression (**Figure 5E**) demonstrated a significant positive relationship between altitude and SD susceptibility score (Y = (3.12 × 10□□)X - 0.625, R^2^ = 0.35, *P* < 0.0001), indicating that higher elevations – including moderate elevations - are associated with increased SD susceptibility. The slope of this relationship (3.12 × 10□□, 95% CI: 2.29 × 10 to 3.95 × 10□□) is consistent with the magnitude and direction of effects observed in each of the individual SD assessment techniques.

## Discussion

Here we report an increased occurrence of migraine aura at moderate to high elevations, drawing from multiple clinical data sources. Additionally, we demonstrate an increased susceptibility to SD in mice exposed to simulated altitudes above sea level. These findings may be of interest for clinical reasons, as an estimated 700 million to 1 billion people worldwide live at or above moderate elevations.^36,37^ Studying migraine in this setting may also help identify candidate mechanisms by isolating responses that mediate the relationship between elevation and aura.

### Migraine aura prevalence at altitude

We show that the prevalence of aura increases with elevation and that this effect includes moderate elevations. Like a study conducted in Peru,^21^ our work supports a relatively monotonic rise in aura prevalence as elevation increases. This contrasts with data from Nepal, where overall migraine prevalence increased at moderate elevations (1,000–2499m) but declined above 2,500m.^20^

It is important to emphasize that increased aura prevalence with altitude conveys potentially different information, with different mechanistic and treatment implications, than a wholesale increase in migraine prevalence. For aura, the known physiological mechanism of SD may serve to narrow the possible mechanisms. As a first step, we can focus on what aspects of the altitude environment can contribute to SD (see below).

### Implications of the different clinical studies

The primary message from the four clinical datasets is that they all converge on increased occurrence of migraine aura with increasing elevation. Despite individual study-specific weaknesses, each Study differs from the other and each dataset was separate and independently collected -- increasing confidence that the convergent signal is indeed real. While the differences across the presented studies limit direct comparison of aura risk, they do provide different angles from which to view the clinical phenomena:

Study 1 provided the least rigorous data, which do not fully account for confounding by site-specific factors (e.g., practice group-based variation in aura recognition and/or documentation/coding), but served to validate our initial clinical observations and justified the more rigorous studies that followed.

Study 2, the two-site cross-sectional comparison between SLC and Cleveland, added rigor by standardizing ascertainment and introducing the VARS to classify aura among participants, which allowed for more directed assessment of aura features, rather than relying on chart documentation. Study 2 also improved on Study 1 by adding participant-level elevation as the exposure rather than relying on clinical site. We observed an increase in the proportion of patients with aura in SLC that was robust to adjusting for other available clinical variables that differed between the two sites (see Supplementary Table S2, Expanded Model).

Study 3, the cross-sectional study of the ARMR tertiary headache clinic database, helped generalize the finding of increased prevalence of aura with altitude (of residence) beyond the anchoring location of SLC. This is important because the phenotype in SLC could possibly have been due to unique features of the location, e.g. a genetic founder effect in the Latter Day Saint population that accounts for ∼50% of the state population. Study 3 also significantly increased the number of subjects examined, and the geographical and altitudinal diversity of that sampling. In Study 3, we went beyond using VARS as a binary classification of aura, using specific aura features within VARS to test their relationships with elevation, and found scotoma alone to be significantly associated with elevation. This finding is of particular interest, since scotoma can retinotopically follow the positive phenomena of aura (scintillations/zig-zag lines), reflecting underlying SD propagation across primary visual cortex.^34^ Scotoma’s apparent association with elevation — in the absence of other independent significant associations with other aura features (in particular, zigzag lines) — suggests that altitude-related physiological changes could preferentially associate with the negative (inhibitory) phase of SD within visual cortex, rather than producing a nonspecific increase in overall aura symptom burden. However tentative, these findings warrant replication in larger samples and are consistent with the hypothesis that hypoxia- and hematologic-driven changes in cortical excitability act on a regionally specific substrate (discussed in more detail in the sections below).

Finally, Study 4, the cross-sectional examination of the Epic Cosmos dataset, expanded our inquiry to the whole continental United States. It confirmed the positive association between migraine aura and altitude, albeit with a different definition of migraine aura. Because of the clinical nature of the dataset, detailed clinical documentation and/or VARS scores were not available, and we used the proportion of ICD-10 migraine with aura (G43.1) to migraine (G43) diagnosis as our proxy for aura prevalence in the sample. Though the definition of aura was less rigorous (dependent on coding and clinician/charting practice-based factors, as was Study 1), the presence of an aura/elevation association in a nationwide sample was arguably the most rigorous confirmation of the phenotype across the studies.

Notably, the estimated effect size of elevation on aura prevalence varied widely across our sub-studies, with a consistent 3.7% and 2.9% increase in aura odds with 100m increased elevation in Study 2 and Study 3 (respectively), contrasting with the much smaller 0.129 to 0.299 percentage point increase in absolute aura prevalence per 100m in Study 4 (a linear regression coefficient, not directly comparable to the odds ratios above). These studies differ considerably in both sampling and participant characteristics (tertiary/academic referral centers vs nationwide sample including both academic and community samples), as well as different definitions of aura (clinician assigned vs. VARS vs. ICD-10). VARS is clearly advantaged by its migraine-specificity and quantitative nature, allowing for a more rigorous definition of aura compared to chart documentation- and/or ICD-10 code-based assessment, which introduces clinician-dependent variability across time and location.^38^ On the other hand, the tertiary headache population represented in Studies 2 and 3 is less generalizable to the population as a whole, because the overall severity and complexity of patient presentation to academic medical centers is higher than in Study 4’s predominantly community based EMR-based sample. Thus, while we may have greater confidence in the sensitivity and specificity of the aura diagnosis in our smaller samples, the larger nationwide confirmation of the significant association between aura and altitude – albeit smaller in (differently measured) effect size – is also persuasive. Ultimately an accurate assessment of the true effect size of elevation on aura prevalence will require larger population-based samples with ascertainment of aura using VARS or other gold-standard equivalents.

### Possible mechanisms of SD susceptibility at altitude

Although migraine prevalence is known to increase with altitude, migraine overall can be influenced by multiple factors, each with multiple potential mechanisms (hypoxia,^9,10^ barometric pressure differences,^11,12^ increased solar radiation/sunlight,^13,14^ metabolic challenge,^15,16^ dehydration,^17^ and a pro-inflammatory environment.^18,19^). In contrast, migraine aura has a known physiological mechanism in SD,^7,8^ which allows us to focus on candidate mechanisms of SD initiation as we speculate about why aura might be more prevalent at altitude. Within this framework, hypoxia and thrombosis/ischemia are plausible because both are essential for brain oxygenation and are well-established triggers or facilitators of SD^7,39,40^.

A potential advantage of an aura phenotype associated with altitude is that it may narrow the parameter space for both human and animal studies. In mice, both SD thresholding and SD counting paradigms showed a graded pattern of SD susceptibility across the three altitudes tested, with the largest effect seen in the high altitude group (4500 m). Although differences at moderate altitude (1400 m) did not reach statistical significance when each assay was analyzed separately, the combined SD susceptibility score highlights the consistent directionality across assays. This suggests that moderate-altitude effects may be modest but aligned with our clinical findings, and useful for guiding mechanistic studies.

#### Hypoxia

Most evidence supporting hypoxia as a key trigger of migraine (and aura) at altitude comes from *acute* oxygen manipulations in humans, particularly normobaric hypoxia studies. In these studies reducing the fraction of inspired oxygen (FiO□) without altering barometric pressure can trigger both migraine aura and migraine headache.^9,10^ However, human physiology studies employing either acute *hypobaric* hypoxia to more closely mimic both the barometric and hypoxic changes of altitude are, to our knowledge, nonexistent.

Acute hypoxia and anoxia are well-established triggers of SD in preclinical studies across multiple species and experimental models.^7^ Severe hypoxia directly stimulates glutamate release from neurons,^41^ and impairs sodium potassium ATPase function in both neurons and astrocytes, disrupting their ability to maintain electrochemical gradients.^42^ Since Na□/K□-ATPase dysfunction, excessive glutamate release, and extracellular K□ accumulation are key drivers of SD initiation,^7^ these findings can support a direct mechanistic link between acute hypoxia and SD susceptibility.

However, the oxygen manipulations typically used to induce acute migraine, aura, or SD, are far greater than the oxygen fluctuations experienced at moderate elevations.^9,10^ In the human studies that induced aura with hypoxia, reducing FiO to ∼12% (from 21% at sea level) resulted in capillary hemoglobin saturations of 70–83%, which is comparable to physiological conditions at altitudes exceeding 4000m. For comparison, effective FiO_2_ at a moderate elevation like SLC (1400m) is 17-18% and capillary hemoglobin saturation is 94-97%.^43^ While these manipulations are certainly relevant to very high-altitude exposure, their implications for more moderate elevations are unclear.

Moreover, to our knowledge there is no physiological data on the *chronic* effects of either normobaric or hypobaric hypoxia on aura or SD susceptibility. This is relevant because individuals residing at altitude undergo physiological adaptations that help maintain adequate tissue oxygenation despite lower oxygen availability.^44,45^

That said, while mild to moderate hypoxia alone may not be sufficient to *directly* trigger SD or aura, it could lower the threshold for SD initiation and/or have indirect metabolic effects (such as a shift toward glycolysis/ lactate production, and oxidative pathway activation^45,46^) that could also make the brain more susceptible to additional triggers. Our mouse findings, using a protocol that simulates chronic altitude exposure^47^, support this concept, aligning with previous work showing that hypoxia exposure—without directly inducing SD—can increase the susceptibility to subsequent SD induction.^48^

#### Thrombosis/Ischemia

In preclinical models, ischemia due to vascular occlusion was one of the first known triggers of SD.^7,40^ Importantly, large vessel occlusions/thromboses are not required to generate SD - models of transient thrombosis/embolism are sufficient to generate SD without apparently leaving permanent damage.^49,50^ SD susceptibility at altitude could thus relate to changes in vascular rheology or thrombotic propensity. Red Blood Cell (RBC) expansion is well-documented at extreme, high, and moderate altitudes.^51,52^ On an anecdotal basis, our writing group has observed relative increases in Hct in SLC migraine patients vs. other clinical settings, though we could not rigorously evaluate this in the current datasets due to missingness. More conclusively, and comparable to prior work,^47^ Hct values measured at the end of the 10-day mouse altitude exposure were modestly but significantly increased in local altitude mice (∼51%) and robustly increased in high-altitude mice (∼70%) compared with sea-level controls (∼49%; **Figure S2D**).

Although RBC increases improve oxygen delivery to tissues, they also elevate blood viscosity and disrupt cerebrovascular perfusion, likely accounting for the increased thrombotic risk at altitude.^53–56^ In a very high altitude population (4300m), migraine (mostly with aura) was significantly more likely in subjects with elevated hematocrit.^57^ There is also evidence that increased RBC mass is correlated with migraine aura in subjects with polycythemia and in subjects with elevated hematocrit in the setting of congenital cardiac disease.^58^

Of course, hypoxia and thrombosis/ischemia are unlikely to be the only altitude-related influences on SD/aura susceptibility. Other factors including increased solar radiation, dehydration, metabolic challenge, and inflammation have all been advanced as independent potential triggers of aura (though definitive evidence remains limited).^59–61^ Moreover, these mechanisms are not always mutually exclusive: metabolic challenge^62^ can amplify hypoxic stress, dehydration^63^ can increase thrombosis risk, and (neurogenic) inflammation (also a known consequence of SD^7,8^) can contribute to a prothrombotic state.^64^ Together, these converging stressors may lower the threshold for SD initiation.

### Limitations and considerations for future work

Our findings pertain to the occurrence of migraine and related aura features in populations that are presumably adapted to their respective elevations. Therefore, while the acute physiology studies in the literature provide valuable insights towards candidate mechanisms, they are not necessarily applicable to our dataset. Our animal work in mice adapted to their respective altitudes begins to address this gap, but further mechanistic work is needed. Second, while our human sample does include individuals residing at high elevations (>2500m), it does not extend to very high elevations (>3500–5500m) so we must exercise caution when comparing our data to studies conducted at higher altitudes. Moreover, the mechanisms driving migraine aura susceptibility at moderate altitudes may differ from those at higher elevations. That said, our mouse model suggests a graded relationship between altitude on SD susceptibility – an effect likely influenced by multiple (and/or overlapping) mechanisms.

Study 1 was designed as a preliminary, hypothesis-generating inquiry in which eligibility required a clinician-assigned migraine diagnosis; patients presenting with only non-migraine headache disorders were therefore not enrolled. As a consequence, other headache subtypes (e.g., tension-type headache, cluster headache) were not systematically tracked or reported. This limits our ability to determine whether the observed difference in aura prevalence between sites reflects an altitude-specific effect on migraine with aura specifically, or a broader difference in headache case-mix between the two tertiary referral populations. This limitation is partially mitigated by the convergence of findings across Studies 2–4, which used more rigorous and standardized inclusion criteria across multiple independent sites and populations.

Another important consideration is that two out of four of the reported clinical studies (Studies 2 and 3) rely on VARS, whereas the others rely on clinical documentation and clinician assigned diagnosis and/or ICD code assignment. Each of the three approaches have their own strengths and weaknesses for aura evaluation: clinical assessment (considered to be the gold standard) is dependent on clinician expertise and completeness of documentation, and ICD-based code assignment is subject to numerous user factors that may impede consistent or accurate assignment of aura-specific sub-codes (favoring assignment of the more simplistic/inclusive parent codes). VARS was developed as a screening and phenotyping tool for aura, and has also been validated for diagnostic classification; while it is not fully reflective of the true diagnostic gold standard of expert-led, clinical diagnosis, it mirrors current ICHD criteria, and was thus selected for the purposes of Studies 2 and 3. Notably, in Study 3, despite the availability of clinician-assigned diagnoses for migraine aura, we could not be confident that clinicians were consistently or specifically assessing for aura, based on the database’s ascertainment methodology.^25,26^ This concern is partially supported by the results of our sensitivity analysis using clinician-assigned diagnoses (Supplementary Table S5), which demonstrated an attenuated association that may stem from variable misclassification of the outcome. Future epidemiologic studies are clearly warranted, and given this data, they will likely need to explicitly consider aura assessment, rather than relying on diagnosis codes.

Our current working model (a hypothesis) proposes that both hypoxia and rheologic changes associated with hematopoietic adaptation to altitude contribute to SD susceptibility, but their relative importance shifts with elevation. At moderate altitudes, rheological factors (increased hematocrit and blood viscosity) may play a dominant role in promoting SD initiation, whereas at higher altitudes, hypoxia and hematologic changes may act synergistically to increase susceptibility. Future studies should attempt to dissect hematologic and hypoxic mechanisms to clarify how altitude-related physiological adaptations influence migraine risk and SD generation across elevations. Now that we have SD as a target mechanism to investigate, and methods exist to separate the elements of the physiological altitude response (e.g. separate manipulation of barometric pressure and oxygen), these questions are now amenable to investigation.

### Clinical implications and conclusions

Globally, between 700 million and 1 billion people dwell above 1000m (3280ft).^37,65^ Assuming (conservatively) global migraine prevalence of 12% and migraine with aura prevalence of 4%,^1,2^ between 28 and 40 million individuals with migraine may be affected by the increased prevalence of aura with altitude (**Figure 6**).

**Figure 6.**
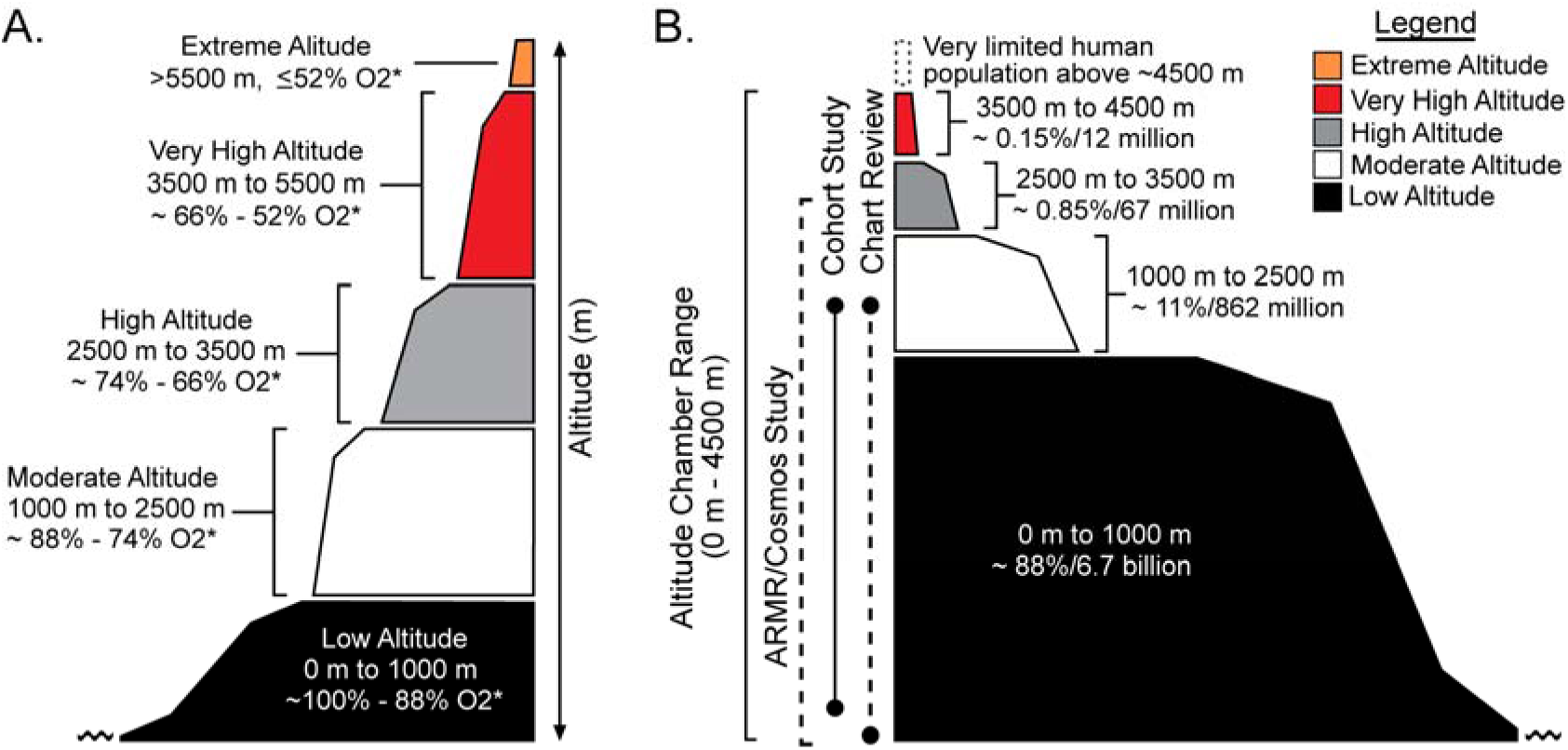
Schematic Illustration of Altitude Zones and their Global Human Populations. **A:** Altitude tier classifications, depicted to illustrate altitude ranges of interest, based on previous literature^1,2^. Shaded areas represent the relative height of each tier (from low altitude, <1000m, to extreme altitude, >5500m). **B:** Human population sizes within each altitude classification tier^3^ (depicted in A; from low altitude to very high altitude). Shaded areas represent a relative, approximate proportion of the total human population living within each tier, extending along the x-axis. The y-axis labels highlight the altitude range of each clinical study and the range our animal study-based altitude chamber. <u>Note</u>: for both A and B, the horizontal axis is arbitrary and does not represent specific distances nor locations. Shading corresponds with altitude tier, as depicted in A and B, summarized in the Legend.

This estimate does not consider (because there is no data upon which to ground it) the possibility that altitude increases the likelihood that individuals with migraine who would otherwise be *without* aura, have aura at altitude. Aura increases the burden of migraine,^4^ and it is also associated with an increased likelihood of stroke and other adverse cardiovascular outcomes.^66,67^ The opportunity to address a mechanistic target (SD) that is also the earliest measurable physiological phenotype of migraine (aura) brings both immediate and potential future benefits. Clinically available drugs could be considered in the current care of higher altitude populations with migraine aura, with the rationale that suppressing an element of the migraine attack generation might have enhanced disease modifying effects compared to other methods. On an investigational level, the mechanistic target represented by SD, along with the known and testable mechanisms involved in the altitude response, could generate drug targets potentially relevant to all migraine with aura.

## Data Availability

All data produced in the present study are available upon reasonable request to the authors

## Acknowledgements

The authors would like to acknowledge Ka-Ho Wong for his assistance with Study 1, Dr. Lindsay Hunter for her assistance with Study 3, and Lauren Pearson DO, MPH for her expertise in hematocrit testing and interpretation.

Research reported in this publication was supported by the National Institute of Neurological Disorders and Stroke of the National Institutes of Health R61 NS 125153 (MMC, KCB), T32NS115723 (KMR), R01 NS 102978, 104742 (KCB), and the Department of Defense Congressionally Designated Medical Research Program (CDMRP) PR20091 (KCB).

## Author contributions

SKB and KCB conceived the study, MMC, KMR, CM, LSM, KCB contributed to the design of the study, MMC, KMR, CM, LSM, JGU, ZAA, AN, JB, KCB acquired and analyzed data, MMC, KMR, CM, LSM, JGU, ZAA, AN, KP, JB, TS, SA, KBD, SKB, KCB were involved in drafting the text or preparing the figures.

## Supplemental Materials

### Supplemental Methods for Human Studies

#### Covariate selection and sensitivity analyses across studies

As the reader may note, the results for all four clinical samples are presented using sex and age as the pre-specified adjustment covariates for the primary regression analyses (presented as the Main Model throughout), treating these as the minimum sufficient set for the relationship between elevation and aura across all four clinical studies. Importantly, both age and sex were available across all studies, while other candidate variables (race, BMI, site, headache days per month, MIDAS, and PHQ-4) were available only in selected studies.

The selection of age and sex as the minimum sufficient coefficient set of interest was also evaluated using a directed acyclic graph (DAG; Figure S1) representing the proposed causal structure among elevation, aura, and the clinical variables available in each dataset (race, BMI, site, headache days per month, MIDAS, and PHQ-4). Using the DAG, we identified four candidate backdoor paths between the exposure (elevation) and outcome (aura): (1) elevation – site – BMI – age – aura; (2) elevation – site – age – aura; (3) elevation – site – race – MIDAS – aura; and (4) elevation – site – race – PHQ-4 – MIDAS – aura. In the proposed DAG, BMI is a collider on path (1), blocking it without adjustment; MIDAS is similarly a collider on paths (3) and (4). To block the remaining open path (2), adjustment for either site or age is sufficient. Because site is strongly collinear with the primary exposure (elevation varies substantially but not exclusively by site in Studies 2 and 3), adjustment for age is preferred, as it captures individual-level variation in the exposure-outcome relationship without absorbing signal attributable to elevation itself. Sex was additionally included given the well-established female predominance of migraine with aura and documented sex and hormonal modulation of SD susceptibility.^1–3^

Beyond their role in blocking open paths, including additional covariates in logistic regression — particularly BMI, MIDAS, and PHQ-4 — risks inducing spurious associations among otherwise unrelated variables through collider stratification bias, a consequence of conditioning on a common effect. Moreover, the addition of non-confounding covariates in logistic regression can paradoxically reduce the precision of the primary exposure estimate due to the noncollapsibility of the odds ratio, an effect that is amplified in moderate-sized samples.^4–6^ Accordingly, race, BMI, site, headache days per month, MIDAS, and PHQ-4 were examined only within designated sensitivity analyses (Expanded Models; Supplementary Tables), in which the direction of the altitude-aura association remained consistent across all specifications, supporting the robustness of the primary finding, as presented in the main body of the manuscript.

### Supplemental Figure for Human Studies

**Figure S1.**
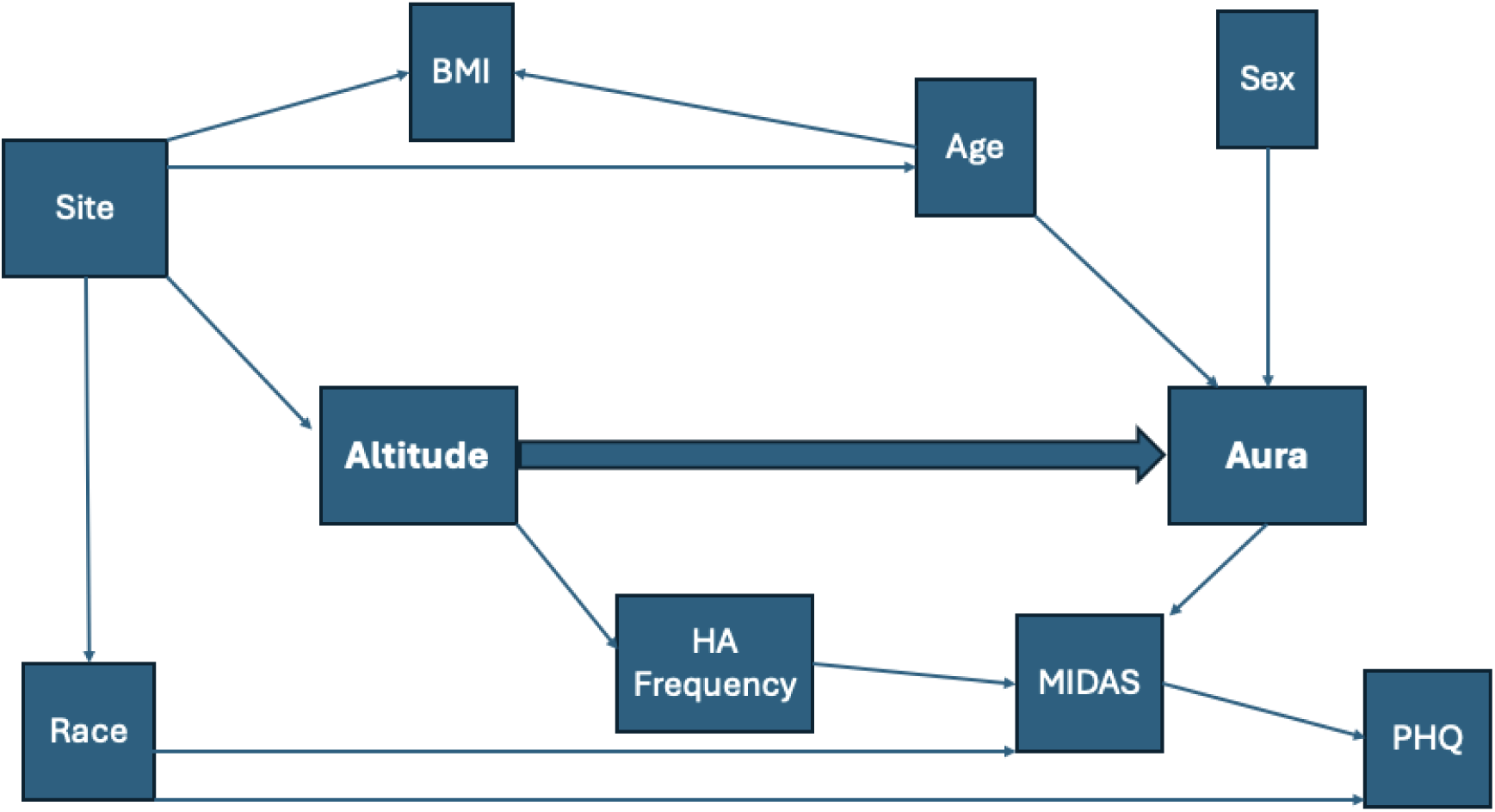
*D*irected acyclic graph (DAG) representing the proposed causal structure among elevation, aura, and the clinical variables available in each dataset (race, BMI, site, headache days per month, MIDAS, and PHQ-4).

### Supplemental Tables for Human Studies

**Table S1:** Logistic regression results for Study 1.

| Model | Term | OR (95%CI) |
| --- | --- | --- |
| Base Model* | Seattle location | 0.187 (0.120, 0.286) |
| Main Model** | Seattle location | 0.182 (0.117, 0.281) |
|  | Age | 0.996 (0.982, 1.011) |
|  | Female sex | 1.271 (0.807, 2.020) |

\*Base Model: site differences only. Reference location, Salt Lake City.
\*\*Main Model: base model, adjusted for age and sex.

**Table S2:**
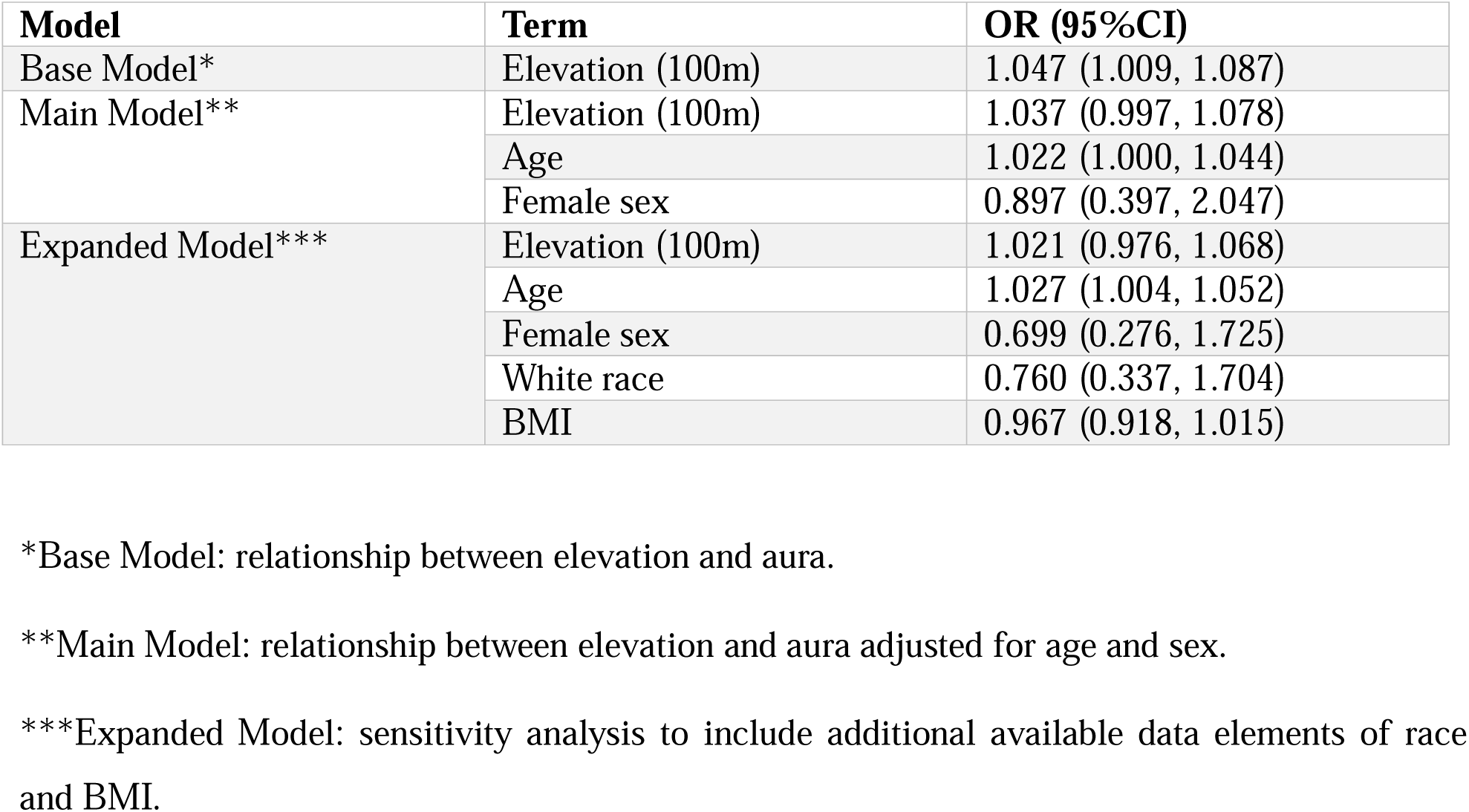
Logistic regression results for Study 2.

**Table S3:** Number of participants per 100m elevation* in Study 3.

| **Elevation category** | **n** |
| --- | --- |
| 0 | 14 |
| 100 | 24 |
| 200 | 55 |
| 300 | 25 |
| 400 | 219 |
| 500 | 26 |
| 600 | 2 |
| 700 | 17 |
| 800 | 30 |
| 900 | 1 |
| 1,000 | 4 |
| 1,100 | 3 |
| 1,200 | 9 |
| 1,300 | 78 |
| 1,400 | 7 |
| 1,500 | 17 |
| 1,600 | 3 |
| 1,700 | 21 |
| 1,800 | 8 |
| 1,900 | 4 |
| 2,000 | 2 |
| 2,100 | 1 |
| 2,200 | 3 |
| 2,300 | 3 |
| 2,400 | 1 |
| 2,600 | 1 |
| 2,700 | 3 |
| 3,000 | 1 |
| 3,200 | 1 |

\* Note: while this table shows the number of participants per 100m elevation category, the primary analysis in Study 3 modeled the relationship between elevation and odds of migraine aura continuously (variable was calculated as participant elevation/100).

**Table S4:** Logistic regression results for Study 3.

| **Model** | **Term** | **OR (95%CI)** |
| --- | --- | --- |
| Base Model* | Elevation (100m) | 1.029 (1.003, 1.056) |
| Main Model** | Elevation (100m) | 1.029 (1.003, 1.056) |
|  | Age | 1.001 (0.990, 1.012) |
|  | Female sex | 1.202 (0.763, 1.948) |
| Expanded Model*** | Elevation (100m) | 1.034 (0.991, 1.078) |
|  | Age | 1.001 (0.986, 1.016) |
|  | Female sex | 0.976 (0.566, 1.736) |
|  | Headache days per month | 1.001 (0.977, 1.026) |
|  | White race | 0.750 (0.262, 2.452) |
|  | Site - Dartmouth | 0.819 (0.504, 1.311) |
|  | Site - Utah | 0.779 (0.420, 1.438) |
|  | MIDAS | 1.005 (1.000, 1.009) |
|  | PHQ-4 | 0.956 (0.892, 1.022) |

\*Base Model: relationship between elevation and aura.
\*\*Main Model: relationship between elevation and aura adjusted for age and sex.
\*\*\*Expanded Model: sensitivity analysis to include additional available data elements of HA days per month, race, MIDAS, PHQ-4 and site. Reference site was Mayo Clinic Arizona.

**Table S5:**
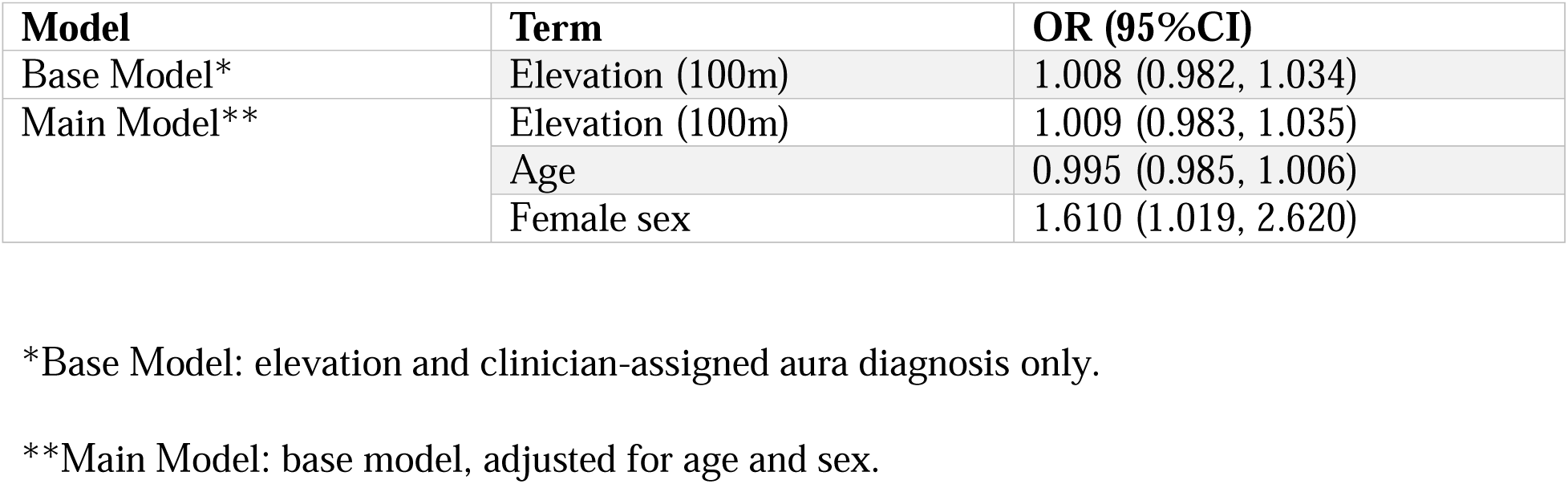
Logistic regression results for Study 3; performed as a sensitivity analysis to evaluate aura as a clinician-assigned ICHD-3 diagnosis. Relative to Table S4, 24 of the initial 884 participants (2.7%) were missing clinician-assigned diagnoses, and therefore excluded.

**Table S6:** Logistic regression results for Study 3 evaluating individual VARS features and elevation. P-values for elevation were obtained using Wald tests and adjusted using the Holm correction for multiple comparisons. Significance at alpha=0.05 is in bold.

| **Individual VARS feature** | **Model** | **Term** | **OR (95% CI)** | **P-value** |
| --- | --- | --- | --- | --- |
| Duration 5-60 minutes | Base Model* | Elevation (100m) | 1.009 (0.984, 1.033) | 0.862 |
|  | Main Model** | Elevation (100m) | 1.009 (0.984, 1.033) | 0.913 |
|  |  | Age | 0.999 (0.989, 1.009) |  |
|  |  | Female sex | 0.840 (0.561, 1.265) |  |
| Gradual onset over at least 5 minutes | Base Model* | Elevation (100m) | 1.026 (0.992, 1.059) | 0.516 |
|  | Main Model** | Elevation (100m) | 1.025 (0.991, 1.059) | 0.564 |
|  |  | Age | 0.987 (0.973, 1.001) |  |
|  |  | Female sex | 0.884 (0.509, 1.624) |  |
| Presence of scotoma (blind spot) | Base Model* | Elevation (100m) | 1.049 (1.022, 1.077) | **0.002** |
|  | Main Model** | Elevation (100m) | 1.052 (1.024, 1.080) | **0.001** |
|  |  | Age | 0.981 (0.970, 0.993) |  |
|  |  | Female sex | 2.401 (1.370, 4.517) |  |
| Presence of zig-zag lines | Base Model* | Elevation (100m) | 1.015 (0.987, 1.042) | 0.852 |
|  | Main Model** | Elevation (100m) | 1.016 (0.989, 1.044) | 0.740 |
|  |  | Age | 1.010 (0.998, 1.021) |  |
|  |  | Female sex | 1.436 (0.886, 2.419) |  |
| Unilateral visual field involvement | Base Model* | Elevation (100m) | 1.013 (0.980, 1.045) | 0.862 |
|  | Main Model** | Elevation (100m) | 1.012 (0.979, 1.045) | 0.913 |
|  |  | Age | 1.001 (0.988, 1.015) |  |
|  |  | Female sex | 0.759 (0.458, 1.307) |  |

\*Base Model: elevation and VARS-based aura diagnosis only.
\*\*Main Model: base model, adjusted for age and sex.

**Table S7:** Linear regression results for Study 4.

| **Model** | **Term** | **Coefficient (95% CI)** |
| --- | --- | --- |
| Base Model* | Intercept | 26.003 (25.750, 26.257) |
|  | County elevation (100m) | 0.299 (0.258, 0.339) |
| Main Model** | Intercept | 30.998 (21.003, 40.993) |
|  | County elevation (100m) | 0.129 (0.014, 0.245) |
|  | Age | −0.035 (−0.133, 0.063) |
|  | Percent female | −0.083 (−0.199, 0.033) |
|  | State cluster |  |
|  | AR | 3.395 (2.768, 4.022) |
|  | AZ | −1.685 (−2.880, −0.490) |
|  | CA | 3.754 (3.216, 4.293) |
|  | CO | 7.460 (5.371, 9.549) |
|  | CT | 8.834 (8.614, 9.054) |
|  | FL | 1.236 (0.897, 1.574) |
|  | GA | 2.610 (2.240, 2.979) |
|  | HI | 3.023 (1.937, 4.109) |
|  | IA | 2.564 (2.193, 2.934) |
|  | ID | 7.556 (6.022, 9.091) |
|  | IL | 2.996 (2.831, 3.161) |
|  | IN | 4.285 (4.037, 4.533) |
|  | KS | 3.480 (3.204, 3.757) |
|  | KY | 5.041 (4.573, 5.508) |
|  | LA | 0.807 (0.425, 1.190) |
|  | MA | 6.880 (6.763, 6.996) |
|  | MD | 1.595 (1.463, 1.726) |
|  | ME | 6.659 (6.258, 7.060) |
|  | MI | 4.903 (4.474, 5.331) |
|  | MN | 7.028 (6.657, 7.400) |
|  | MO | 3.588 (3.464, 3.713) |
|  | MS | 2.626 (2.388, 2.864) |
|  | MT | 5.849 (4.652, 7.045) |
|  | NC | 2.113 (2.017, 2.210) |
|  | ND | 5.296 (4.806, 5.786) |
|  | NE | 4.009 (3.461, 4.557) |
|  | NH | 7.125 (6.889, 7.360) |
|  | NJ | 4.104 (3.924, 4.285) |
|  | NM | 3.115 (1.245, 4.986) |
|  | NV | 8.335 (6.624, 10.046) |
|  | NY | 4.151 (3.925, 4.376) |
|  | OH | 8.052 (7.767, 8.336) |
|  | OK | 2.405 (2.192, 2.619) |
|  | OR | 8.508 (7.870, 9.146) |
|  | PA | 5.244 (4.988, 5.499) |
|  | RI | 4.488 (4.361, 4.616) |
|  | SC | 1.791 (1.617, 1.965) |
|  | SD | 4.240 (3.625, 4.856) |
|  | TN | 2.762 (2.593, 2.931) |
|  | TX | 2.532 (2.130, 2.934) |
|  | UT | 8.734 (6.849, 10.620) |
|  | VA | 4.249 (4.047, 4.450) |
|  | VT | 6.546 (6.202, 6.890) |
|  | WA | 5.911 (5.441, 6.381) |
|  | WI | 7.234 (6.890, 7.579) |
|  | WV | -4.426 (-4.815, -4.038) |
|  | WY | 5.948 (4.141, 7.756) |

\*Base Model: county elevation and electronic-medical record-based aura diagnosis only.
\*\*Main Model: base model, adjusted for age, county percent female, and state.

### Supplemental Methods for Animal Studies

#### Mice

A total of 144 C57Bl/6J mice were used across all experiments, including 30 for pilot studies, 72 for SD thresholding, and 42 for SD counting. Of the 72 mice used in SD thresholding studies, 6 mice (*n* = 2 per altitude) were excluded before analysis and unblinding. The most common exclusion reasons were inadvertent SD triggering during preparation; one mouse was excluded due to bleeding during surgical preparation. In SD counting studies, 42 mice were tested, with two exclusions: one from lack of SD induction, one from camera failure preventing SD counting.

#### Surgical preparation

Experiments were conducted in each mouse under isoflurane anesthesia. Following anesthesia induction (3 – 4% isoflurane), mice were positioned in a stereotaxic frame assembly and respiration rate was maintained at 75-85 breaths/min throughout the experiment (1.1-1.8% isoflurane). Body temperature was monitored at 15-minute intervals throughout the experiment (Acorn Series thermometer, Oakton Instruments) and maintained at 36.5 – 37.5°C using a heating pad warmed via a circulating water bath (TE-8D, Techne). After head-fixation in the stereotaxic frame, the skull was exposed and small diameter craniotomies/burr holes were carefully drilled using a pneumatic dental drill (Henry Schein Inc., Melville, NY, USA). For SD thresholding experiments, two burr holes (∼0.5mm diameter) were made in the right hemisphere for SD initiation (AP: +3.0mm ML: +1.0mm) and detection (AP: - 3.0mm, ML: +3.0mm). For SD susceptibility experiments, we followed our previously published methods with minor surgical modifications.30–32 Briefly, a ∼1.5mm diameter craniotomy was created in the right barrel cortex (AP: −1.5mm ML: +2.5mm) to expose the brain for SD initiation (see SD Methods below).

#### Electrophysiology

Extracellular recordings were used to detect SD in thresholding experiments. A borosilicate glass micropipette (1-3.5 MΩ) containing a Ag/AgCl wire was filled with normal saline (0.9% NaCl) and carefully placed in the cortex through the caudal burr hole (∼200μm depth from the skull surface) for recording of local field potentials (LFP). Recordings were referenced to a Ag/AgCl ground wire carefully inserted into the contralateral cortex via an additional burr hole created in the left hemisphere. Signals were amplified at 10kHz using a Multiclamp 700B amplifier, A/D-converted using a Digidata 1332 digitizer and displayed in Clampex 9.2 software (Molecular Devices, LLC., San Jose, CA, USA). After electrode insertion, a 30 – 45-minute baseline was recorded to ensure stable anesthesia (based on respiratory rate and cortical LFP activity^7,8^). LFP recordings then served to confirm SD in real-time via the emergence of the large characteristic direct current shift (DC shift) in the extracellular slow potential (<1Hz) and subsequent suppression of spontaneous cortical activity (1-70Hz).

#### SD thresholding

SD threshold was evaluated using our previously published methods, with minor modifications.30 Briefly, focal 1M KCl was applied into the rostral burr hole via a large diameter (∼22-24µm) borosilicate glass pipette (Sutter Instruments) connected to a pneumatic pico-pump. KCl volume was stepwise increased every five minutes using 20ms square-wave pressure pulses (increasing by 2.5 psi up to 15 psi) until SD was detected via the emergence of the DC shift at the distant LFP electrode. The distant locations for SD initiation and detection (∼6mm) ensured unambiguous confirmation of SD, such that only KCl volumes capable of inducing SD propagation across the entire ipsilateral hemisphere were detected (i.e., no partial SDs). Each pressure ejection was visually monitored using a dissection microscope. The KCl pipette was promptly removed from the burr hole immediately after the delivery of KCl and throughout the 5-minute interval to prevent excessive KCl application due to capillary action.

#### SD counting

SD counting procedures were performed using our previously published methods, with minor modifications. The entire dorsal surface of the skull was illuminated using a white light-emitting diode and intrinsic optical signals (IOS) were collected (0.5Hz) using a CCD Camera (Mightex). Mineral oil was utilized to enhance skull translucency, facilitating optical access. A small barrier composed of cyanoacrylate glue was created around the craniotomy ( to prevent mineral oil from encountering the exposed cortex. This barrier also served to restrict the application of perfusion solutions to the site designated for SD induction. After surgical procedures, saline (0.9%) was slowly (0.016mL/min) perfused onto the cortex via a syringe pump connected to a blunt 34-gauge MicroFil flexible needle (World Precision Instruments). During this 15-minute baseline period, no SDs were detected in any mice. After baseline imaging, saline was replaced with a 1M KCl solution that was perfused for a total of 2 hours (0.016mL/min). Off-line analysis (prior to unblinding) included plotting the z-axis profile of image sequences in Fiji software^9^ to count SD totals over the 2-hour period.

#### SD susceptibility score calculation

In addition to analyzing SD thresholding and SD counting data separately, a post hoc z-score normalization approach was used to generate an exploratory, integrative “SD susceptibility” score combining data from both types of experiments. Because the two datasets differ in scale and units, each dataset was standardized independently by transforming values into z-scores reflecting deviation from the mean of that dataset. For SD thresholding data, the reciprocal (1/x) of the observed SD threshold (in psi) was computed prior to z-scoring so that higher values consistently reflected greater SD susceptibility across both experimental techniques. This composite metric was not pre-specified and is not the primary outcome measure.

#### Statistics, Reporting of Data, Rigor

Sample size determination and power analyses were calculated using G*Power 3.1 software^10^. A pilot feasibility study in male C57Bl/6J mice (n = 30; 10.4±0.4 weeks of age; 10/group) was used to establish the timeline of altitude exposures and inform sample size planning for SD thresholding studies. Based on group mean SD thresholds from the pilot (0m: 7.0; 1400m: 6.8; 4500m: 4.3 in psi; see SD thresholding methods) and the median within-group standard deviation (± 2.7psi), we estimated an omnibus one-way ANOVA effect size of f = 0.45. Using α = 0.05, f = 0.45, and target power ≥ 90%, the required total sample size was n = 66 mice (22 mice/group), which was therefore set as the target sample size for SD thresholding experiments.

Sample size for SD counting experiments was calculated based on published work from our lab where we found a significant difference in SD count in mice using this method (two sample comparison, Cohen’s d = 1.52 for data shown in Figure 6).24 Using α = 0.05 and assuming that the largest contrast would be between 0m and 4500m, power analysis indicated that 11 mice/group would provide ≥ 90% power for this comparison.

Data in Results were plotted and statistical analyses were conducted using Graphpad Prism 10.1.1 (GraphPad Software, LLC, San Diego, CA). Grubbs’ (α = 0.05). D’Agostino and Pearson tests were used to identify outliers and determine if data were normally distributed, respectively. Statistical significance for normally distributed data (Hematocrit data and SD counting data) was evaluated using Ordinary one-way ANOVA with Bonferroni’s multiple comparisons test. For nonparametric data (SD thresholding data), the Kruskal-Wallis test with Dunn’s multiple comparisons test was used to evaluate statistical significance. Effect sizes were calculated from the raw data as Cohen’s d, with 95% confidence intervals estimated by bootstrap resampling, and are reported alongside other statistics.

### Limitations of Animal Studies

Sex differences in SD susceptibility have been reported previously in mice, including lower SD thresholds in females compared to males^2^ as well as modulation of SD susceptibility by ovarian hormones^11^ SD threshold has also been shown to vary across the natural estrous cycle in female mice, with cycle stage–dependent differences in susceptibility^12^.

For the present study, we therefore chose to use male mice to minimize hormonal variability in SD susceptibility that could confound interpretation of altitude-related effects. Accurate estrous-cycle staging requires repeated sampling over multiple consecutive days (Cora, M.C. et al., *Toxicologic Pathology,* 2015) which was not feasible in the current experimental design because mice remained continuously at their assigned simulated altitude until the day of testing. Daily removal of female mice from altitude chambers for cycle determination would have introduced additional fluctuations in hypoxic exposure and atmospheric pressure, constituting a significant experimental confound.

One important limitation is the use of anesthesia, as anesthetics can modify SD thresholds and susceptibility. Isoflurane anesthesia dose-dependently suppresses SD *in vivo* ^13,14^ and *in vitro* ^15^, likely through multiple mechanisms. These may include combinatory effects such as reducing cerebral metabolism (via decreased cerebral metabolic rate of oxygen (CMRO_2_)^15^, attenuating glutamatergic transmission (through antagonism at the glycine site of NMDA receptors)^16^, enhancing GABAergic transmission^17^, and increasing CBF (presumed vasodilation)^18^. As such, in our study we may underestimate altitude effects on SD thresholds and/or susceptibility.

### Supplemental Figures for Animal Studies

**Figure S2.**
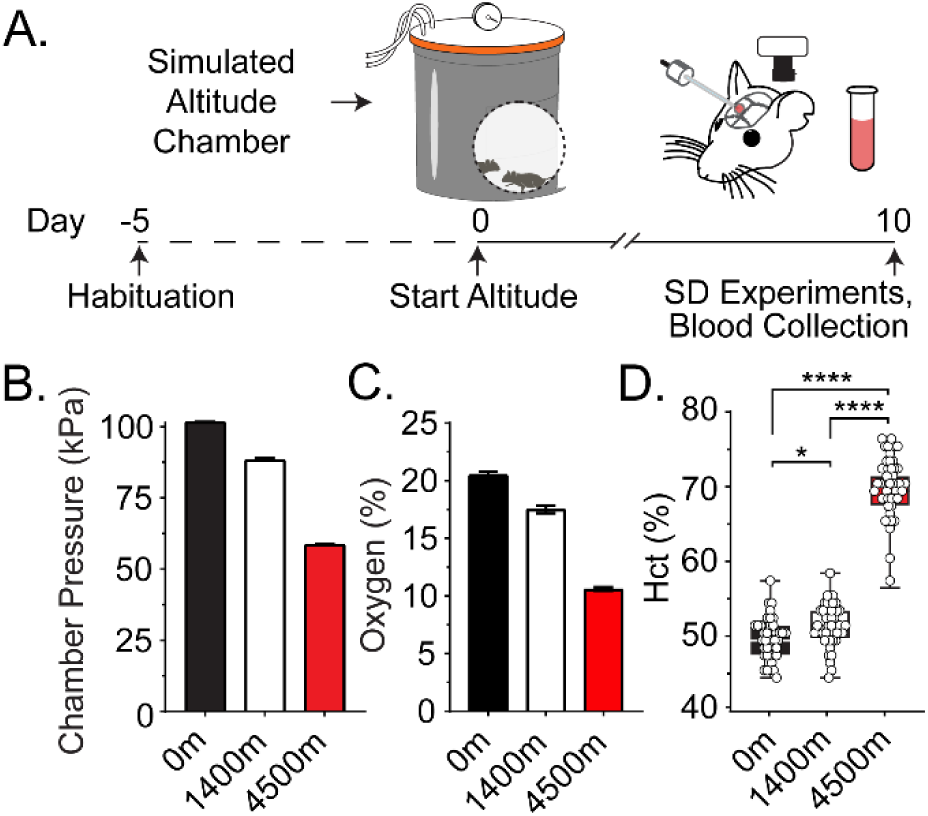
Experimental timeline and altitude exposure. **A.** Experimental timeline of study. Mice were transferred from standard mouse cages into custom-made altitude chambers and habituated (dashed line) for 2-5 days prior to altitude manipulations (day 0). On the morning of day 10, mice were gradually returned to local lab conditions for experiments assessing SD threshold and susceptibility. Top schematic shows the design of the custom-made altitude chambers, followed by blood collection and SD experiments. **B:** Average atmospheric pressure sensor data from 0m (101.6 ± 0.1 kPa, n = 32 chambers), 1400m (88.5 ± 0.4 kPa, n = 32), and 4500m (58.6 ± 0.2, n = 34) altitude chambers. **C:** Sensor data showing the effective concentration of oxygen (%) inside 0m (20.4 ± 0.3%, n = 14 chambers), 1400m (17.4 ± 0.3%, n = 5 chambers), and 4500m (10.5 ± 0.2, n = 18 chambers) altitude chambers. **D:** After SD experiments (data shown in Figure 5), blood was collected, and Hct was measured. Group Hct data shows that Hct is increased in mice exposed to 4500m (*n* = 46 mice) compared to 1400m (*n* = 44) and 0m (*n* = 41) altitudes. \*\*\*\**P* < 0.0001, \**P* = 0.013; One-way ANOVA with Bonferroni’s post hoc multiple comparisons test.

